# A Multilevel Framework for Evaluating the Impact of Monoclonal Antibodies Against Respiratory Syncytial Virus in Daycare and Households

**DOI:** 10.64898/2026.11.03.26367194

**Authors:** Ke Li, Virginia E. Pitzer, Daniel M. Weinberger

## Abstract

Monoclonal antibodies (mAbs) reduce the risk of severe disease caused by respiratory syncytial virus (RSV) among immunized infants. The potential of these products to indirectly protect unimmunized contacts remains incompletely understood. This uncertainty is, in part, due to our limited understanding of how mAbs may influence population-level RSV transmission. Here, we develop a multiscale simulation framework that integrates RSV transmission dynamics across households and daycare centers to explore the characteristics of the population that could influence the strength of indirect effects of infant mAb immunization. The framework incorporates individual-level heterogeneity in age and time-varying infectiousness, as well as demographic variation in household composition and contact patterns, to capture realistic transmission pathways across close-contact networks. Using a virtual population informed by U.S. household characteristics, we show that infant immunization would only be expected to provide modest indirect protection to toddlers, and the magnitude of these effects depends on contact structure and intervention coverage. Our findings demonstrate the importance of integrating fine-scale transmission networks into evaluations of RSV prevention strategies and provide a generalizable framework for assessing population-level impacts of interventions against respiratory pathogens.

## Introduction

Respiratory syncytial virus (RSV) is a leading cause of infant hospitalization. Evidence has shown that long-acting monoclonal antibodies (mAbs) are effective at preventing severe disease in newborns [1–8]. However, the question of whether this protection provides indirect benefits for unimmunized household members or daycare contacts has not been evaluated. It has generally been assumed that monoclonal antibodies, which provide a relatively short duration of protection in the first year of life, would not have a major impact on transmission, and therefore there would not be indirect protection. However, in settings with clustering of infants, like daycare centers or households, there is a possibility that mAbs could have some effect on disrupting transmission if they prevent asymptomatic infection as well as severe disease.

Addressing these gaps requires a model that explicitly represents both the biological mechanism of protection of mAbs on infection and the contact structure of the population. Prior modeling work has established that household transmission is a major contributor to infant RSV infection, with siblings thought to be the primary source of infection [9,10]. Household composition and size substantially influence estimated maternal RSV vaccine impact [11,12]. However, existing frameworks have examined household transmission separately from other settings, such as daycare centers, where transmission events occur more frequently among children [13,14]. In addition, no models have coupled these settings with an explicit, individual-level natural history model capable of distinguishing between symptomatic and asymptomatic infectiousness [15].

Here, we develop a multiscale agent-based stochastic model integrating daycare, household, and community contacts while explicitly tracking each individual’s infection history, including infection-blocking mAbs, asymptomatic breakthrough infections, and transmission chains. We use this framework to quantify direct and indirect protection from infant mAb immunization across different coverage levels, identify the transmission pathways underlying indirect effects, and evaluate their sensitivity to daycare mixing and household composition.

## Results

### Model Simulation Framework

To investigate the direct and indirect protective effects of infant mAb immunization against RSV infection, we first developed an agent-based stochastic transmission model that links three nested environments of exposure: daycares, households, and the external community (**Fig. 1A**). In daycares, infants, toddlers, and staff mix directly. These individuals are generated from the household population and coupled to daycares through daily attendance. Infections acquired at daycare can be carried home, and household-acquired infections can, in turn, be introduced into the daycare setting. External community risk represents importation of infection from the community, which is independent of either setting. Within the population structure, each individual’s infection history follows a natural-history model (**Fig. 1B**). Unprotected individuals progress through a reinfection framework, in which each following infection confers partial immunity that reduces susceptibility to subsequent episodes [16]. mAb-protected infants follow a different pathway: they experience an initial period of complete protection that wanes into a phase of standard susceptibility, during which they can have breakthrough infections (*I*_*ab*_) or wane into *S*1 if they are not exposed to RSV. We assume that these breakthrough infections are asymptomatic and less infectious (i.e., 20% as infectious as *I*1, see **Supplementary Materials**). If they are infected while still protected from symptomatic disease by mAbs (*M*_*I*_), we assume that they receive partial immunity against future reinfection, eventually waning into the *S*2 state. We fitted the model to recent time-varying mAb effectiveness data [1] to calibrate the duration of immune protection parameters (see **Methods** and **Fig. S1**). To characterize population-level variability, we simulated *N* independent cohorts, each with a stochastically generated population and household composition, while applying the same transmission structure and individual disease process (**Fig. 1C**).

**Figure 1.**
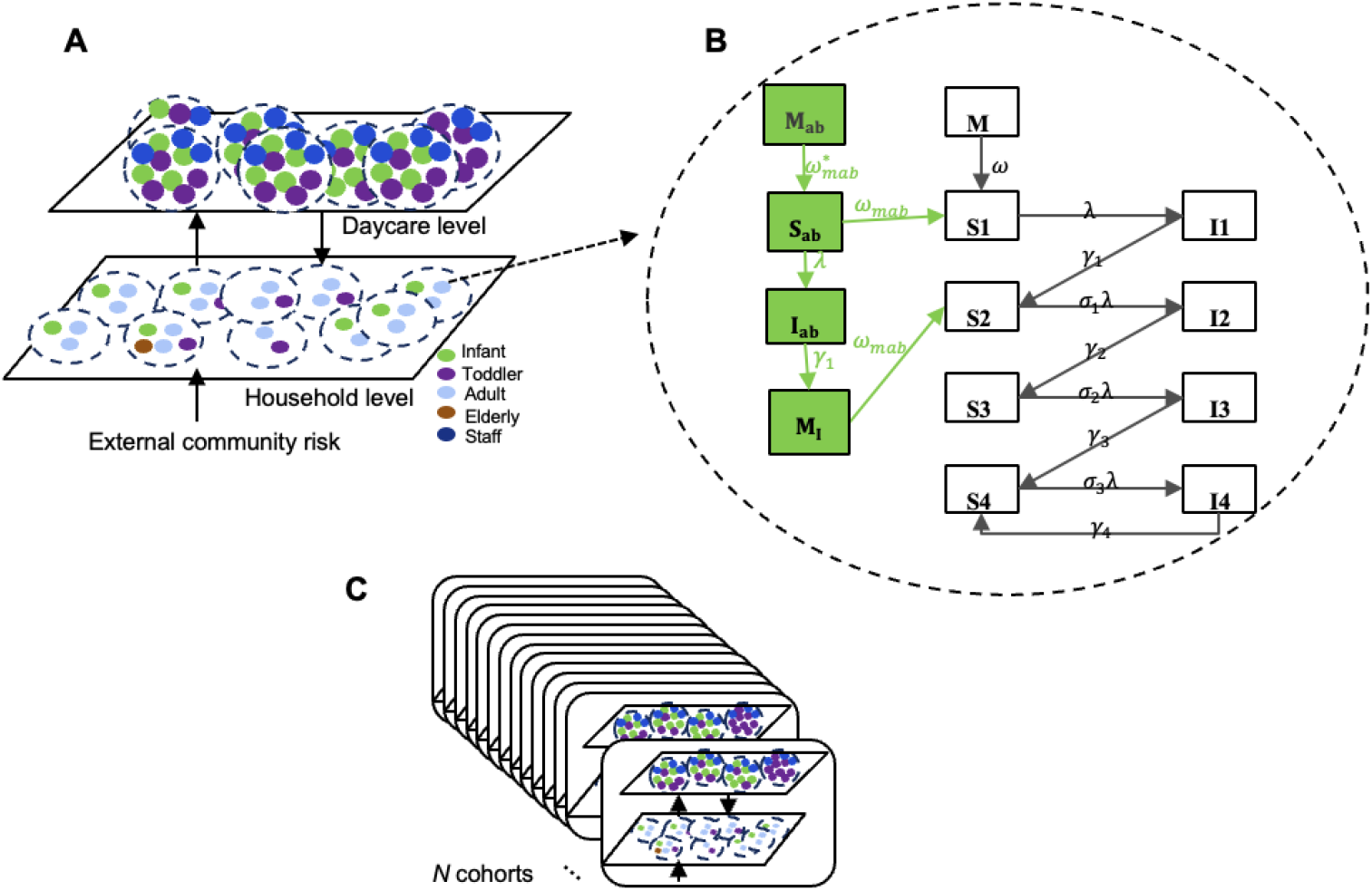
Multiscale structure of the agent-based RSV transmission model. **(A)** Individuals are organized across daycare classrooms and households, coupled by daily attendance, plus an external community risk term. Colors represent different roles. **(B)** Individual-level infection history for one representative agent: unprotected individuals (white box) progress through a reinfection process with decreased susceptibility (σ_1−3_′ 0 < σ_3_ < σ_2_ < σ_1_ < 1) after each episode; mAb-protected infants (green boxes) follow a parallel pathway in which breakthrough infection during the susceptible window is assumed to be asymptomatic (*I*_*ab*_) but remains infectious and provides partial immunity against future reinfection once immunity from mAb wanes. **(C)** The complete model structure is independently replicated to generate *N* simulated cohorts.

### Daycare and household structure

We simulated 50 independent cohorts (see **Table S1** for parameter values). The resulting daycare and household structures are summarized in **Table 1** and reproduce key features of realistic daycare settings while introducing variability across simulated cohorts. Classrooms contained an average of 8.7 children, with the majority (69.2%) classified as mixed-age rooms containing both infants (<1 year old) and toddlers younger than 18 months, while the remaining 30.8% consisted of older toddlers (18 months–4 years). This structure reflects the model’s design to capture age-specific contact patterns by allowing close-contact mixing between infants and young toddlers while separating older toddlers into single-age rooms.

**Table 1.** Daycare and household structure. Each simulated daycare center enrolled a fixed 90 infants (<1 year old) and 90 toddlers (1-4 years old), and children-staff ratio is fixed at 3. The remaining, variable structural characteristics are summarised below (mean (n) and proportion (%) across 50 cohorts). Details of household composition are provided in **Table S2**.

| Characteristic | Value |
|---|---|
| **Daycare structure** |  |
| Mean classroom size, children | 8.7 |
| Mixed-age (young) classrooms, n (%)<sup>a</sup> | 14.4 (69.2%) |
| Single-age (older) classrooms, n (%)<sup>b</sup> | 6.4 (30.8%) |
| **Household structure** |  |
| Mean Households per daycare center | 140.0 (129.0-153.0)<sup>c</sup> |
| Infant + toddler siblings, n (%) | 25.6 (18.5%) |
| Infant only, n (%) | 64.4 (45.9%) |
| Toddler only, n (%) | 50.0 (35.6%) |
| Households with two caregivers, % | 85.4 (75.9-95.1) |
| Households with a grandparent, % | 7.8 (2.8-12.7) |

<sup>a</sup> Mixed-age (young) classrooms contain both infants and toddlers younger than 18 months.
<sup>b</sup> Single-age (older) classrooms contain only toddlers aged 18 months or older.
<sup>c</sup> range

At the household level, each cohort comprised an average of 140 households. Household structures were designed to reflect key demographic characteristics of U.S. families, including the predominance of two-caregiver households and limited multigenerational co-residence observed in national surveys [17]. Only 18.5% of the households contained both an infant and a toddler, while 45.9% contained an infant without a toddler and 35.6% contained a toddler without an infant. Most households (85.4%) included two caregivers (e.g., parents), and a smaller proportion (7.8%) included a co-resident grandparent.

### RSV epidemics and mAb protection

We then simulated RSV epidemics across increasing levels of infant mAb coverage and calculated corresponding protection effects (**Fig. 2**). Here, we assumed that infections in the *I*_*ab*_ compartment represented asymptomatic infections that would not contribute to detected RSV cases. Under this assumption, we observed a reduction in detected infant infections with increasing mAb coverage, with seasonal peak infections declining from approximately 250 cases at 0% coverage to approximately 100 cases under 100% coverage (**Fig. 2A**). In contrast, we observed minimal changes in RSV prevalence among toddlers (**Fig. 2B**) and adults/older adults (**Fig. 2C**) across coverage scenarios, with epidemic trajectories largely overlapping across all six coverage levels.

**Figure 2.**
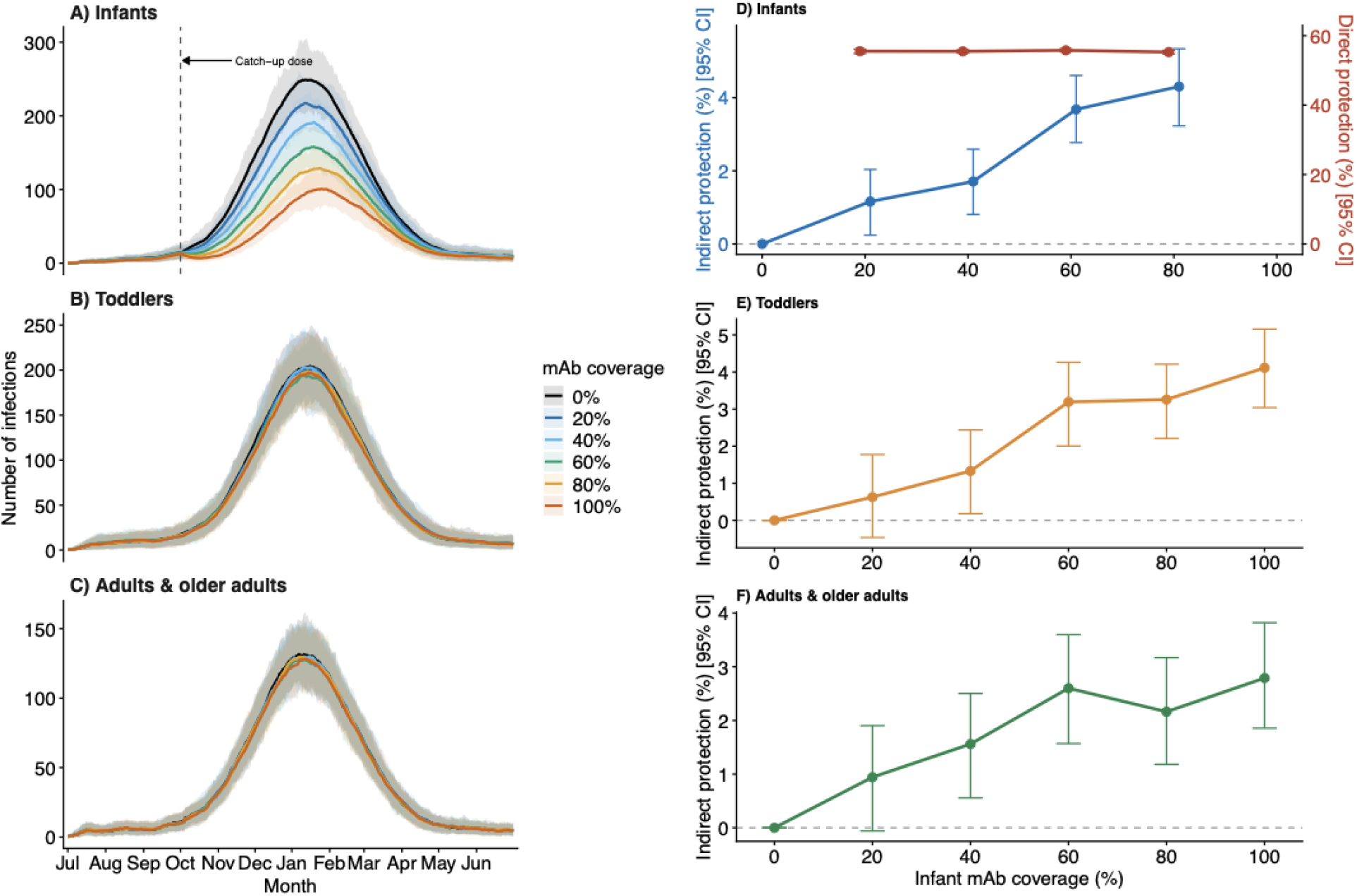
Simulated RSV incidence and protection against infection across infant mAb coverage levels. Mean daily number of RSV infections (shaded ribbons, 95% simulation interval) among **(A)** infants **(B)** toddlers, and **(C)** adults & older adults across six infant mAb coverage scenarios. An RSV season is defined as the period from July of the previous year through June of the following year. The dashed vertical line marks the timing of catch-up dosing and the start of the RSV epidemic. Protection (lines, %; vertical error bars, 95% CI) as a function of infant mAb coverage: **(D)** direct and indirect protection among infants who received mAbs, **(E)** indirect protection among toddlers, and **(F)** indirect protection among adults/older adults. Results are aggregated across all 50 simulated cohorts. Daycare staff were excluded from the calculation.

We next quantified direct and indirect protection for different age groups (see **Methods**). We found that direct protection among infants remained stable across coverage levels, consistent with our validation (**Fig. 2D** and **Fig. S1**). We observed comparatively modest indirect protection among unimmunized contacts. Indirect protection increased gradually with coverage, reaching approximately 4.3% (95% CI: 3.3–5.4%) among infants at 80% coverage (**Fig. 2D**) and 4.1% (95% CI: 3.0–5.3%) among toddlers at 100% coverage (**Fig. 2E**). We also observed a similar but smaller increase among adults and older adults, reaching approximately 3% protection at full coverage (**Fig. 2F**). We also calculated the direct and indirect protection against hospitalization and found that the indirect protection was even smaller for hospitalization than for infection, with only a small effect among infants and toddlers, and no detectable effect among adults (**Fig. S2**). We further explored an alternative scenario in which *I*_*ab*_ infections were assumed to be mildly symptomatic and detectable; under this assumption, the reduction in detected infant cases was substantially attenuated, with no large decline in infant infections observed, and other quantities remain the same (**Fig. S3**).

### Transmission changes under mAb coverage

To identify which transmission pathways were affected by infant mAb coverage, we compared the number of transmission events between 0% and 100% coverage, separately for daycare and household settings (**Fig. 3**). In daycare, we observed that toddler-toddler and toddler-infant transmission accounted for the majority of transmission events across both coverage levels. We also observed the largest relative reductions among pathways involving infants: infant-infant transmission decreased by 46% (1.64 to 0.88 events), and infant-toddler transmission decreased by 54% (4.86 to 2.22 events), consistent with reduced infant transmission under mAb protection. In contrast, transmission pathways that did not directly involve infants (e.g., staff-infant, staff-toddler, and toddler-toddler) showed comparatively small reductions (2–13%).

**Figure 3.**
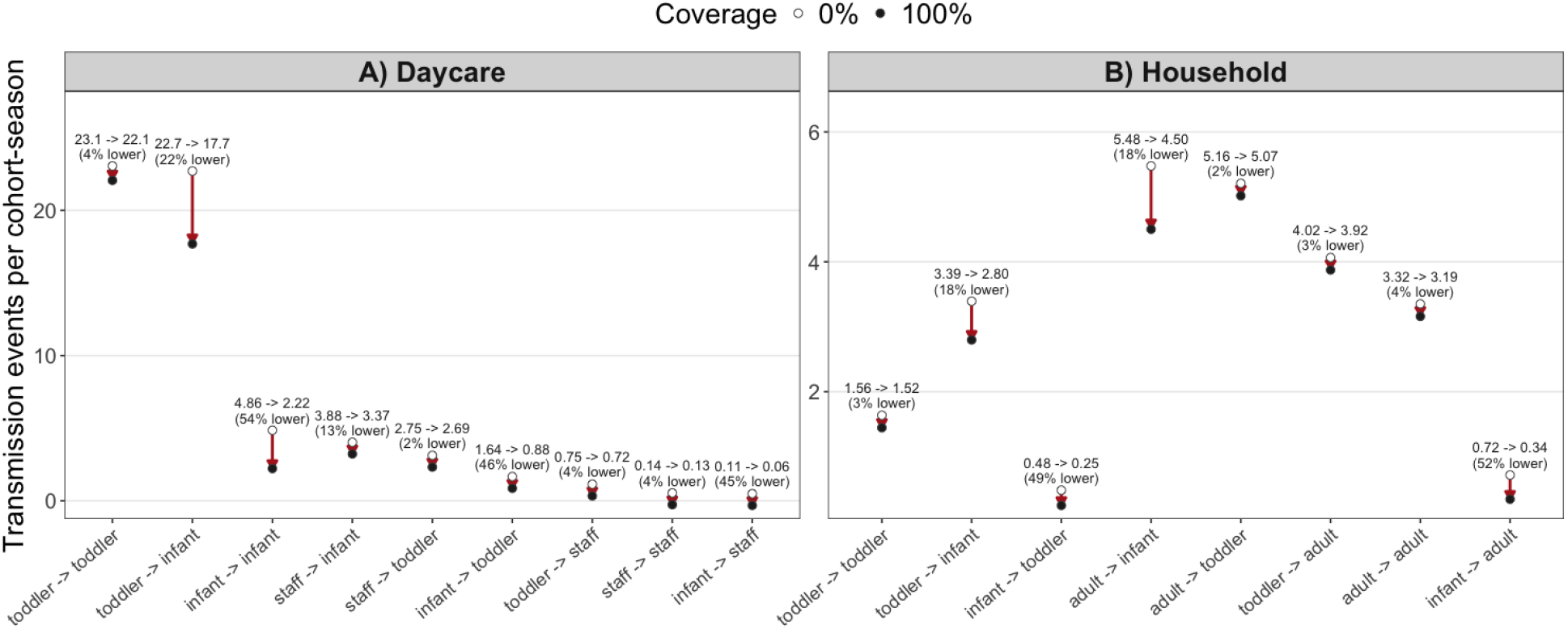
Transmission pathways by setting and mAb coverage. Mean transmission events per cohort-season for each infector-infectee pathway at 0% (open circles) and 100% (filled circles) infant mAb coverage, within **(A)** daycare and **(B)** household settings. Pathways are ranked by event count at 0% coverage within each panel. Labels show raw event counts at each coverage level and the percentage reduction from 0% to 100% coverage. Note that the adult group shown in the figure combines adults and older adults.

In households, adult-associated transmission pathways, including those involving older adults, accounted for the majority of overall transmission. Adult-infant transmission represented the largest single pathway (5.45 events at 0% coverage), while toddler-adult, adult-adult, and adult-toddler pathways each contributed similarly high numbers of transmission events (3.1–5.2 events). Similar to the daycare setting, we observed the largest relative reductions for pathways originating from infants, with infant-adult and infant-toddler transmission decreasing by 52% (0.72 to 0.34 events) and 49% (0.48 to 0.25 events) under full mAb coverage, respectively. However, these represent a small proportion of overall transmission events in households. In contrast, pathways not involving infants showed minimal changes.

### Structural assumptions and indirect protection

To evaluate whether the limited indirect protection observed above resulted from specific structural assumptions rather than a fundamental limitation of infant mAb intervention, we repeated the analysis under two alternative daycare and household structures. In Scenario 1, we relaxed the assumption that cross-room transmission in daycare occurs only through staff, allowing all individuals (i.e., infants, toddlers, and staff) to travel between classrooms. In Scenario 2, we increased the proportion of infant-anchored households containing a co-resident toddler sibling, thereby reducing the frequency of infant-only households in the simulated population structure (**Table S3**).

Direct protection among immunized infants remained unchanged under both alternative structures, consistent with our validation against the mAb effectiveness data (**Fig. 4A** and **4B**). In contrast, we observed greater indirect protection under both alternative scenarios. Indirect protection at 100% coverage increased to approximately 9% among toddlers and 6.5% among adults and older adults in Scenario 1, and increased to approximately 10% and 9%, respectively, in Scenario 2 (**Fig. 4C** and **4D**). However, we found that indirect protection among infants did not change compared with the baseline structure. This is because the number of infant-infant contacts did not change substantially between the primary and the scenario analyses.

**Figure 4.**
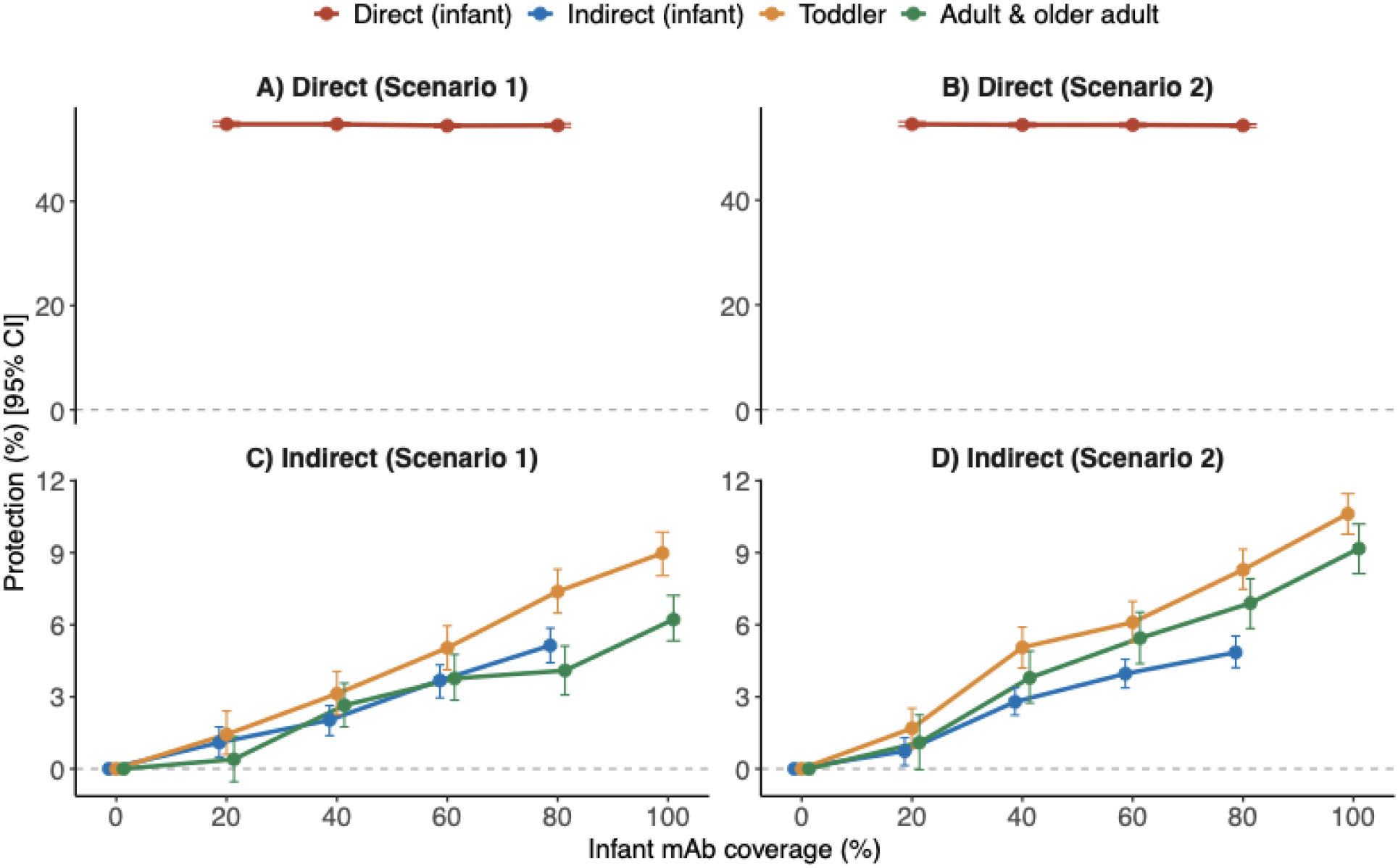
Sensitivity of direct and indirect protection against infection to daycare and household structural assumptions. Estimated protection (lines, %; vertical error bars, 95% CI) as a function of infant mAb coverage under two alternative model structures. Scenario 1: cross-room daycare transmission permitted for all attendees, not only staff. Scenario 2: increased proportion of infant-anchored households containing a co-resident toddler sibling. **(A, B)** Direct protection among vaccinated infants. **(C, D)** Indirect protection among infants, toddlers, adults, and older adults.

We further compared pathway-specific transmissions between the two alternative scenarios and the base case in daycare and household settings (**Fig. S4-S5**). In daycare, we found that both scenarios increased infant-associated transmission relative to the base case. Toddler-infant transmission became the dominant pathway in daycare under both scenarios and the dominant pathway in households under Scenario 2 (**Fig. S4**). These pathways also showed larger reductions with increasing coverage. In contrast, pathways not involving infants showed minimal changes, indicating that the enhanced indirect effects were driven primarily by infant-linked transmission. In households, we observed that Scenario 1 had a limited effect on pathway magnitudes, whereas Scenario 2 substantially increased toddler-infant transmission, from 5.15 events in the base case to 16.1 events per season at 0% coverage (**Fig. S5**). This pathway also showed a larger reduction with increasing coverage, demonstrating that increased infant–toddler co-housing enhanced the potential for indirect protection. We also evaluated a scenario in which *I*_*ab*_ infections remained fully infectious. We found that indirect protection among infants was reduced to approximately zero (**Fig. S6**).

## Discussion

In this work, we developed a multiscale agent-based mathematical framework integrating daycare, household, and community transmission to evaluate how mAb immunization provides direct and indirect protection against RSV infection for different age groups. We found that indirect protection among unimmunized contacts was modest under realistic U.S. demographic and household structures, with toddlers driving most daycare transmission and adults accounting for the majority of household transmission. However, indirect protection was sensitive to variation in daycare and household structures. We showed that increasing contacts between infants and other age groups, or modifying household composition (e.g., increasing infant–toddler co-residence), could increase observed indirect protection. The findings provide insight into potential study designs to measure indirect protection.

Previous modeling studies have suggested that infant RSV mAb could generate indirect benefits by reducing RSV transmission [18]. They demonstrated that the estimates depend on assumptions regarding contact patterns and the relationship between prophylaxis, infection, and infectiousness. However, the model did not capture detailed age- and setting-specific contacts or the distinct roles of daycare, household, and community transmission, nor did it explicitly represent the stages of protection, infection, and infectiousness following mAb administration. These simplifications may limit its ability to identify the transmission pathways driving indirect protection and quantify how these benefits vary across age groups. Therefore, we need to construct models that explicitly account for household and daycare transmission to quantify the level of indirect protection across different contact environments. In particular, the magnitude of indirect protection can depend on how immunized infants are embedded within age-structured contact networks, including their interactions with high-transmitting groups in settings such as daycares and households. Our findings identify the conditions under which passive mAbs can generate indirect protection and support a cautious interpretation of their population-level impact: although indirect protection is biologically plausible, it is modest relative to direct protection and depends on the underlying transmission structure and contact patterns. In particular for RSV, infants accounted for relatively few transmission events in both daycares and households, which explain why the indirect protection is weak.

In simulations using defined population structure, we found that household transmission was primarily driven by adults in the U.S. population structure. This result contrasts with a recent U.S. household transmission study, which showed that the median age of index cases was 6 years, with most index cases aged 6 months to 12 years[19]. The difference is because we did not include a school-age contact role (e.g., 5–17 years) in the household structure, limiting our ability to capture the transmission mediated by school-age children observed in the studies. Other observational studies have similarly highlighted the importance of sibling-dominated transmission, with older siblings serving as a key source of infection for younger children within households [20–23]. This difference was driven by household composition, as most infants in our simulated population did not have a co-resident toddler sibling. When we assumed a higher proportion of infant–toddler pairs within households (e.g., Scenario 2), transmission shifted toward being largely toddler-driven. This result also highlights that intrinsic infectiousness alone does not determine realized transmission. We have shown previously that children have higher RSV infectiousness than adults and older adults, as indicated by higher peak viral loads, longer shedding, and higher predicted transmission probabilities [24].

We demonstrate that indirect protection is not simply correlated to direct protection, but is sensitive to the specific structure of daycares or households. Our scenario analyses showed that indirect protection could be twice as high depending on contact assumptions, either by allowing broader daycare mixing or by increasing infant–toddler co-residence in the household. Together, these results indicate that the generalizability of indirect protection estimates depends on the household or daycare contact structures that shape transmission. Without accounting for these structural differences, comparisons across studies, countries, or populations may misattribute effects of demographic composition and social mixing to underlying biological mechanisms. Future studies designed to quantify indirect effects should therefore explicitly consider population-specific contact structures.

We also provide a generalizable framework for evaluating targeted immunization strategies against other pathogens beyond RSV. By explicitly separating intervention-specific biological characteristics (e.g., effectiveness and duration of protection) from population-level structural features (e.g., contact patterns, age mixing, and household composition), this framework enables evaluation of how indirect benefits emerge from interactions between intervention mechanisms and transmission. This principle is evident across diverse pathogens. When the targeted age group constitutes a primary reservoir driving transmission to the broader population, indirect protection among unvaccinated individuals can be substantial [25,26]. By contrast, in closed communities, vaccinating a specific age group has produced strong herd protection for others [27], whereas similar strategies in more open community settings have resulted in modest or undetectable indirect effects [28–30]. Similarly, a targeted household-based immunization strategy supported by strong theoretical rationale demonstrated limited real-world indirect benefit, likely because vaccination did not fully cover the actual contact network surrounding the protected individual [31]. Together, these examples imply that the population-level impact of an intervention depends not only on its biological efficacy but also on the position of the targeted group within the underlying transmission network. This has important study design implications. The indirect protection expected under certain study designs may be stronger (or weaker) than the indirect protection expected under real-world conditions. For example, a study of the indirect effects of mAbs restricted to households with both an infant and a toddler could overestimate indirect protection relative to the average U.S. household composition.

Some limitations should be considered when interpreting our results. First, community risk was modeled as a fixed, coverage-independent term, which likely understates any feedback between reduced local transmission and reduced reintroduction risk from the community. This is a simplification that could lead the indirect effect to understate the benefit achievable at high coverage, since a lower-transmission population would also import fewer cases from outside. Second, we did not explicitly include school-age children and instead used school-based contact patterns to represent daycare contacts, as our focus was on daycare, and no daycare-specific contact patterns were available. In addition, the structural scenarios examined here represent only a small subset of real-world childcare arrangements and household compositions. Populations with different classroom sizes, staff-to-child ratios, and multigenerational living patterns may exhibit indirect effects outside the range reported here. Broader exploration of this structural parameter space is needed to establish the generalizability of these estimates across settings.

In summary, our study suggests that mAbs are only expected to provide weak indirect protection, even in settings with substantial mixing between infants and other unimmunized individuals, such as households and daycare, and the strength of observed effect can depend on population contact structure. The framework offers a generalizable approach for evaluating targeted immunization strategies across diverse populations.

## Methods

### Model overview

We developed a stochastic, individual-based transmission model coupling daycare centers, households, and community exposure over a single simulated RSV season. See full model details and parameter values in the **Supplementary Information**.

### Agent-based transmission model

Each agent’s infection history was simulated using a compartmental model that tracked transitions among maternally protected, monoclonal antibody (mAb)-protected, and unprotected states. These transitions were represented using Eqs. (1)-(4) in **Text S1** and **S2**.

### Force of infection

The daily probability that a susceptible individual *i* at time *t* becomes infected is modelled as π_*i,t*_ = *P*(infection *i* at *t* | susceptible *i* at *t*–1) = 1 − exp(–ϕ_*R*(*i*)_ λ_*i,t*_), where ϕ_*R*(*i*)_ is role-specific susceptibility, and λ_*i,t*_ is the total force of infection, aggregated across the three environments, such that λ_*i,t*_ = λ_*i,t*_ ^*comm*^ + β_*HH*_ λ_*i,t*_ ^*HH*^ + β_*DC*_ λ_*i,t*_ ^*DC*^, where

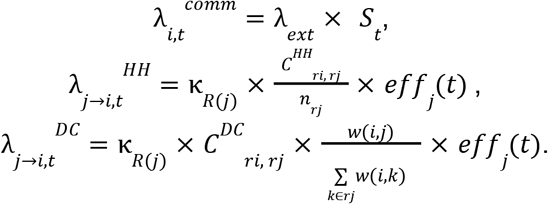

Here, λ_*ext*_ is baseline community transmission rate, and *S*_*t*_ is a time-varying seasonal forcing term. The seasonal component was informed by national surveillance data in the US in the 2024/25 season. κ_*R*(*j*)_ is the role-specific infectivity, and *eff*_*j*_ (*t*) is the time-varying, viral-load-derived infectiousness [24] of infectious contact *j* (Eqs. (5)-(7) in **Text S3**). *C*^*HH*^ and *C*^*DC*^ are role-structured contact matrices from [32], and *w*(*i, j*) is a same-/different-classroom contact weight. Full models and their descriptions are provided in Eqs. (8)-(14), **Text S4** and **S5**.

### Direct, total, and indirect protection

For coverage level *c*, let *AR* denote the attack rate in group *g*, then

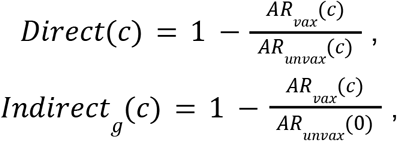

where group *g* ∈ {infant, toddler, adult and elderly}. We combined adults and older adults when calculating indirect protection. 95% confidence intervals were obtained by nonparametric bootstrap (2000 samples) over simulation replicates (Eqs. (15)-(16), **Text S6**).

### Calibration of mAb duration of protection

We calibrated two parameters governing the duration of mAb-conferred protection (the mean length of the fully-protected phase, ω*_*mab*_ , and the subsequent waning phase, ω_*mab*_) to empirical effectiveness data from [1]. Using an age-structured compartmental model driven by the same seasonal force of infection, we reproduced the cross-sectional cohort design. We then fit the two parameters by minimizing the least-squares difference between the model-predicted and observed month-by-month effectiveness over the first year following mAb administration (Eqs. (17)-(32), **Text S7**).

## Data Availability

All data produced in the present work are contained in the manuscript

## Acknowledgments

This work was supported by the National Institute of Allergy and Infectious Diseases of the National Institutes of Health under Award Number K99AI199123 awarded to K.L. This work was also supported by a grant from the National Institutes of Health (R01AI137093) awarded to D.M.W and V.E.P. The content is solely the responsibility of the authors and does not necessarily represent the official views of the National Institutes of Health.

## Code Availability

We used the R statistical software (v4.3.2) for all analyses and visualization. The code used in this study is publicly available on Github: https://github.com/keli5734/RSV_hh_daycare.

