## supplementary figures and texts for "A Multilevel Framework for Evaluating the Impact of Monoclonal Antibodies Against Respiratory Syncytial Virus in Daycare and Households": Supplementary Figures.pdf

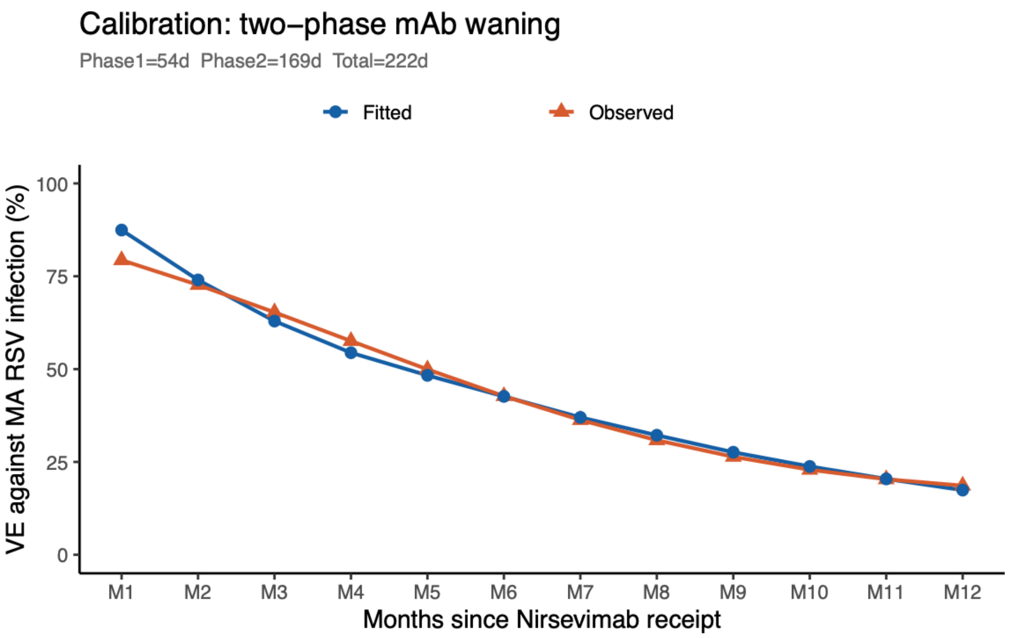


**Figure S1. Calibration of nirsevimab-mediated protection.** Model-fitted (blue circles) and observed (orange triangles) effectiveness against RSV infection following nirsevimab administration are compared over 12 months after receipt.

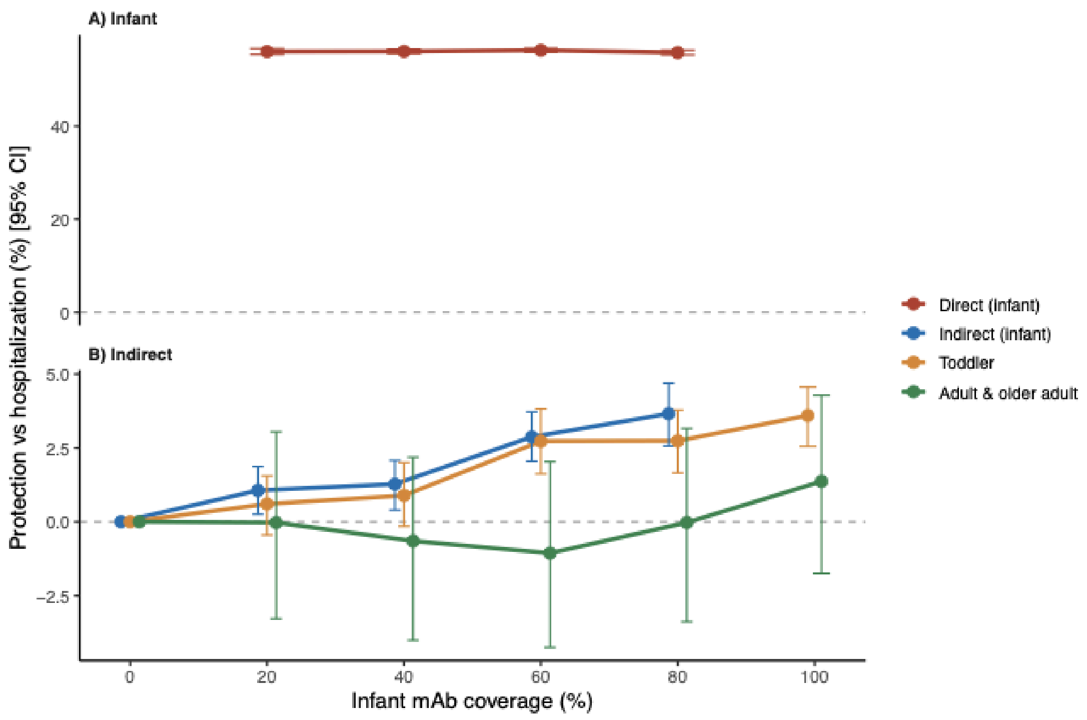


**Figure S2. Protection against hospitalization across infant mAb coverage levels.** Protection (lines, %; vertical error bars, 95% CI) as a function of infant mAb coverage: **(A)** direct protection among infants who received mAbs, **(B)** indirect protection among infants, toddlers and adults/older adults. Results are aggregated across all 50 simulated cohorts.

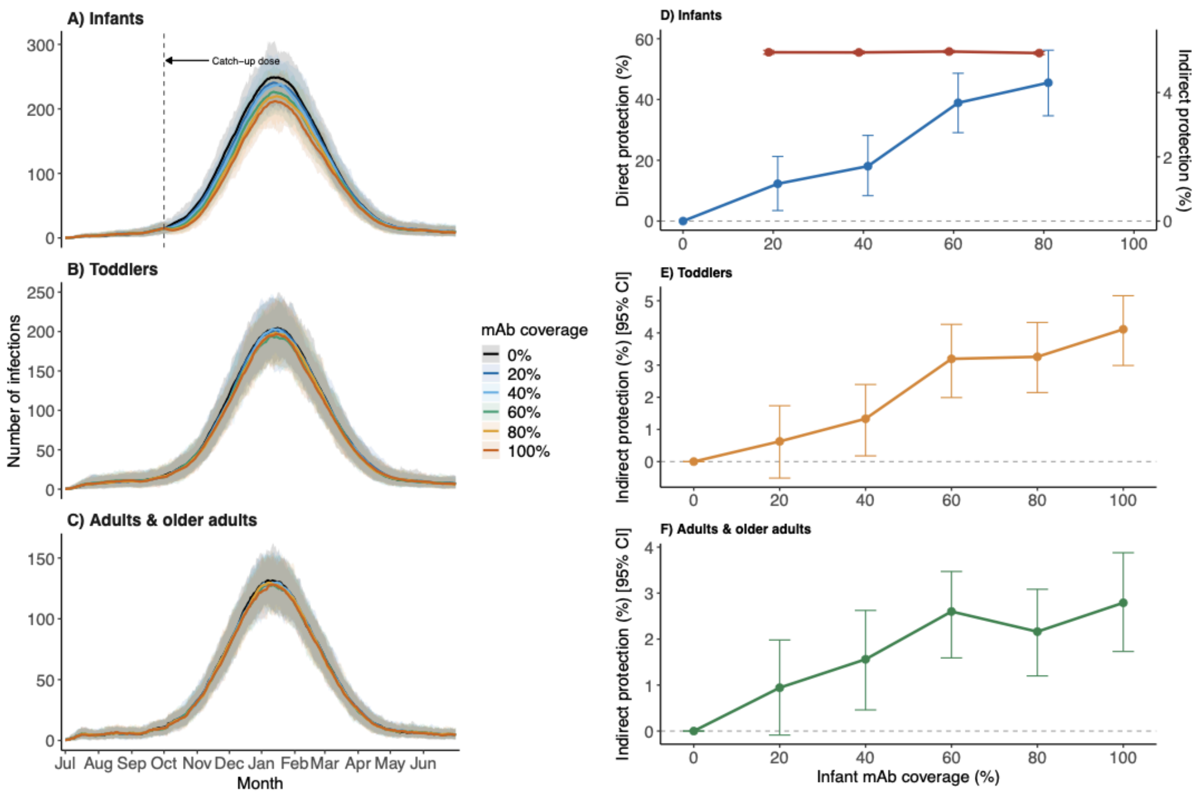


**Figure S3. Simulated RSV incidence and protection across infant mAb coverage levels.** Mean daily number of RSV infections (shaded ribbons, 95% simulation interval) among **(A)** infants **(B)** toddlers, and **(C)** adults & older adults across six infant mAb coverage scenarios. The dashed vertical line marks the timing of catch-up dosing and the start of the RSV epidemic. Estimated protection (lines, %; vertical error bars, 95% CI) as a function of infant mAb coverage: **(D)** direct (red curve) and indirect (blue curve) protection among infants who received mAbs, **(E)** indirect protection among toddlers, and **(F)** adults/older adults.

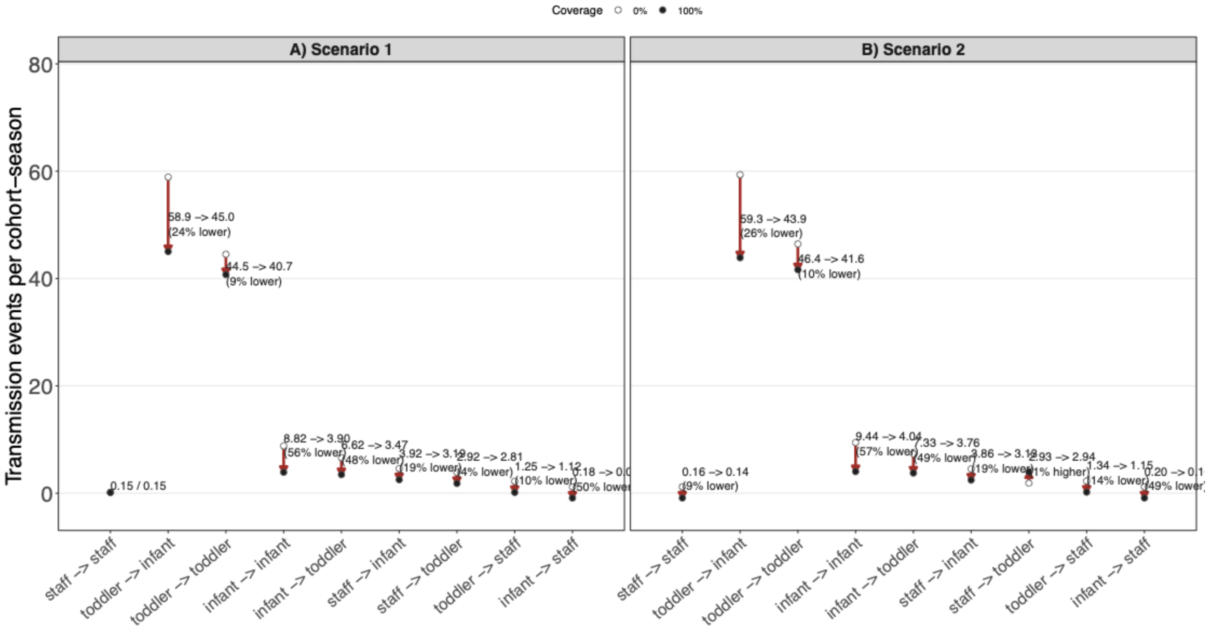


**Figure S4. Transmission pathways in daycares in alternative assumptions.** Mean transmission events per cohort-season for each infector-infectee pathway at 0% (open circles) and 100% (filled circles) infant mAb coverage, under **(A)** scenario 1 and **(B)** scenario 2. Labels show raw event counts at each coverage level and the percentage reduction from 0% to 100% coverage.

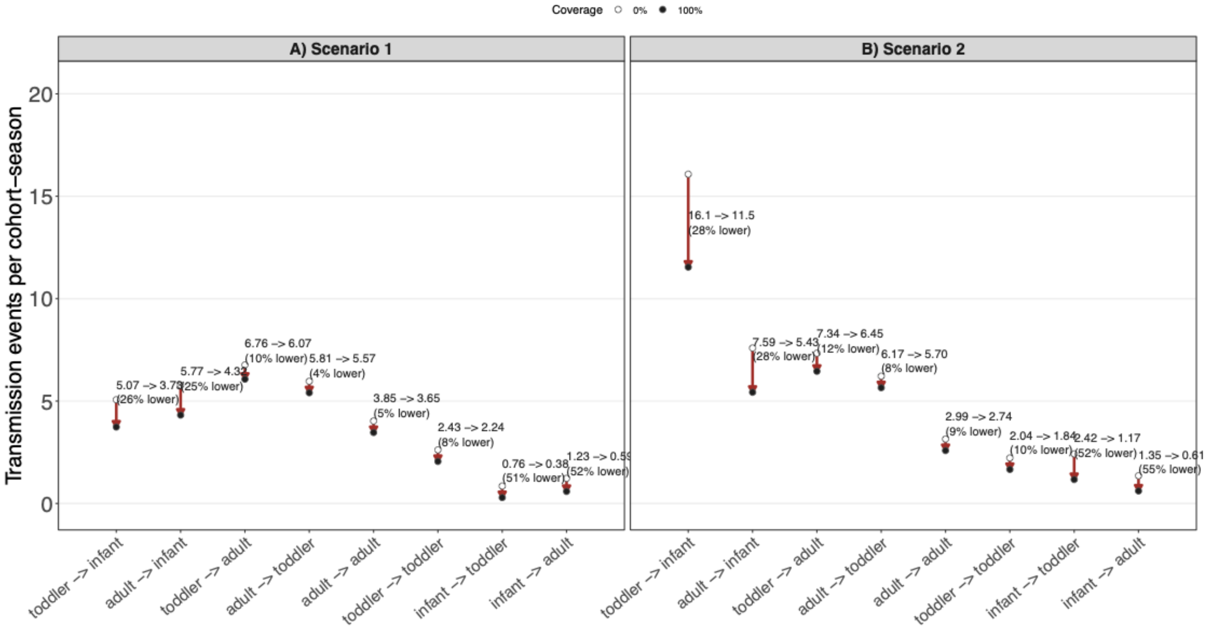


**Figure S5. Transmission pathways in households in alternative assumptions.** Mean transmission events per cohort-season for each infector-infectee pathway at 0% (open circles) and 100% (filled circles) infant mAb coverage, under **(A)** scenario 1 and **(B)** scenario 2. Labels show raw event counts at each coverage level and the percentage reduction from 0% to 100% coverage.

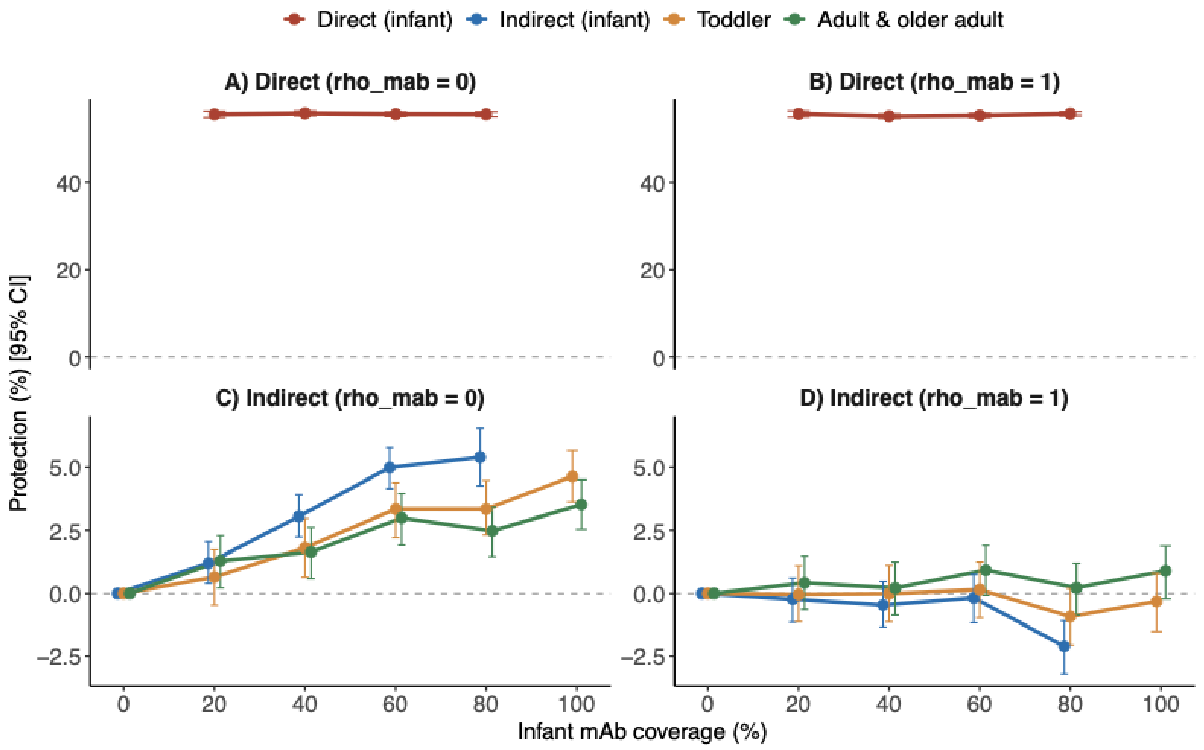


**Figure S6. Protection across infant mAb coverage levels under alternative assumptions for the relative infectiousness of breakthrough infections while protected by mAbs.** Protection (lines, %; vertical error bars, 95% CI) as a function of infant mAb coverage: **(A, B)** direct protection among infants who received mAbs, **(C, D)** indirect protection among infants, toddlers, adults/older adults. Results are aggregated across all 50 simulated cohorts.

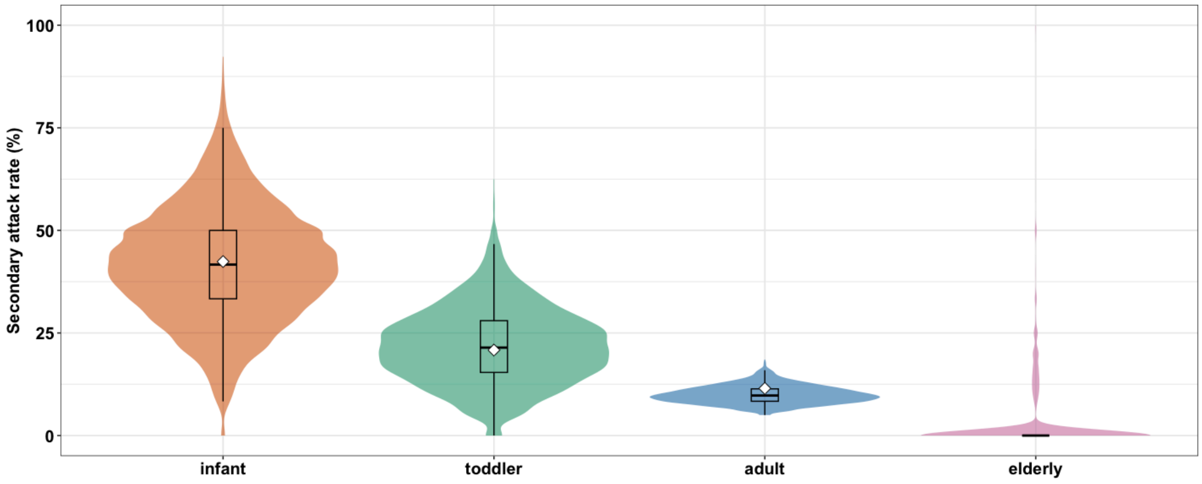


**Figure S7. Secondary attack rates by age group.** Violin plots show the distribution of secondary attack rates (SARs) of RSV among infants, toddlers, adults, and older adults across simulated households. White diamonds indicate the observed data used for calibration, and boxplots show the median and interquartile range. Note that we did not calibrate SARs to data for the elderly group.
