## supplementary figures and texts for "A Multilevel Framework for Evaluating the Impact of Monoclonal Antibodies Against Respiratory Syncytial Virus in Daycare and Households": Supplementary Text and Table.pdf

### Supplementary Materials

#### Notation

$i, j$: individuals; $R(i) \in \{\text{infant, toddler, adult, elderly}\}$; $t$: simulation day; $n_i(t)$: cumulative prior infections; $\tau$: days since an individual's own infection onset.

#### S1. Infection status transitions

State transitions occur with daily probability $1 - e^{-\omega \Delta t}$ ($\Delta t = 1$ day):

$$M \xrightarrow{\omega_M} S, \qquad M_b \xrightarrow{\omega^*_{mab}} S_{ab}, \qquad S_{ab} \xrightarrow{\omega_{mab}} S, \qquad MI_{ab} \xrightarrow{\omega_{mab}} S, \tag{1}$$

$$\omega_M = \frac{1}{112}\ \text{d}^{-1}, \qquad \omega^*_{mab} = \frac{1}{53.6}\ \text{d}^{-1}, \qquad \omega_{mab} = \frac{1}{168.6}\ \text{d}^{-1}. \tag{2}$$

Infection maps $S \to I$ (symptomatic) or $S_{ab} \to I_{ab}$ (breakthrough infection); recovery maps $I \to S$ or $I_{ab} \to MI_{ab}$, at day $t + T_i$, where infectious duration is

$$T_i \sim \text{Gamma}\left(\text{shape} = 3,\ \text{scale} = \frac{\mu(n_i)}{3}\right), \qquad \mu(n_i) = \begin{cases} 10 & n_i = 1 \\ 7 & n_i = 2 \\ 5 & n_i \geq 3. \end{cases} \tag{3}$$

Relative susceptibility and infectiousness as a function of episode count:

$$\sigma_n = \begin{cases} 1 & n = 0 \\ 0.76 & n = 1 \\ 0.60 & n = 2 \\ 0.40 & n \geq 3, \end{cases} \qquad \rho_n = \begin{cases} 1 & n = 1 \\ 0.75 & n = 2 \\ 0.51 & n \geq 3. \end{cases} \tag{4}$$

### S3. Viral load and infectiousness

$$C_t(\tau, R(j)) = \begin{cases} C_{peak}(R(j)) + r(R(j))\,[t_{peak}(R(j)) - \tau] & \tau \leq t_{peak}(R(j)) \\ C_{peak}(R(j)) + d(R(j))\,[\tau - t_{peak}(R(j))] & \tau > t_{peak}(R(j))\,, \end{cases} \qquad \tau < 0 \Rightarrow C_t = Ct_{LOD}. \tag{5}$$

$$f(C_t) = \left[1 + \exp\left(\frac{C_t - \mu_{vl}}{s_{vl}}\right)\right]^{-1}, \qquad \mu_{vl} = 32.79, \quad s_{vl} = 1.92. \tag{6}$$

Effective infectiousness of an infectious individual $j$ at time $t$:

$$\text{eff}_j(t) = \kappa_{R(j)}\, \rho_{n_j(t)}\, [\beta_1 + \beta_2\, f(C_t(\tau_j, R(j)))] \times \begin{cases} \rho_{mab} & \text{state}_j(t) = I_{ab} \\ 1 & \text{otherwise.} \end{cases} \tag{7}$$

### S4. Force of infection, by setting

**Community (Phase 0).**

$$\lambda_i^{\text{comm}}(t) = \lambda_{ext} \times \frac{S_t}{\max_t\, S_t}, \tag{8}$$

where $S_t$ is the seasonal forcing term.

**Household (Phase 1).** For household $h \ni i$ and $n_{R(j)}^{(h)}$ the count of role-$R(j)$ members in $h$:

$$\lambda_i^{HH,1}(t) = \sum_{\substack{j \in h \\ j \neq i}} C_{R(i),R(j)}^{HH}\, \frac{\text{eff}_j(t-1)}{n_{R(j)}^{(h)}}. \tag{9}$$

**Daycare (Phase 2).** Let $P(t)$ be the set of present individuals; for $i \in P(t)$ in classroom $c(i)$:

$$\lambda_i^{DC}(t) = \sum_J C_{R(i),J}^{DC}\, \frac{w_{in} \displaystyle\sum_{\substack{j \in P(t) \cap J \\ c(j) = c(i)}} \text{eff}_j(t) \;+\; w_{bw}(R(i), J) \displaystyle\sum_{\substack{j \in P(t) \cap J \\ c(j) \neq c(i)}} \text{eff}_j(t)}{w_{in}\, n_i^{J,same}(t) + w_{bw}(R(i), J)\, n_i^{J,diff}(t)}, \tag{10}$$

$$w_{bw}(R(i), J) = \begin{cases} w_{bw,staff} & R(i) = \text{staff or } J = \text{staff} \\ w_{bw,child} & \text{otherwise.} \end{cases} \tag{11}$$

(In the base-case parameterization, $w_{in} = 1$, $w_{bw,staff} = 1$, $w_{bw,child} = 0$, restricting cross-classroom transmission to staff-mediated contact.)

**Household (Phase 3).** For children newly infected at daycare on day $t$ (set $D(t)$), using immediate ($\tau = 0$) infectiousness:

$$\text{eff}_j^{bh}(t) = \kappa_{R(j)}\,\rho_{n_j(t)}\,\left[\beta_1 + \beta_2 f(C_t(0, R(j)))\right] \times \begin{cases} \rho_{mab} & \text{state}_j(t) = I_{ab} \\ 1 & \text{otherwise,} \end{cases} \qquad j \in D(t), \tag{12}$$

$$\lambda_i^{HH,2}(t) = \sum_{\substack{j \in h \\ j \in D(t)}} C^{HH}_{R(i),R(j)} \frac{\text{eff}_j^{bh}(t)}{n^{(h)}_{R(j)}}. \tag{13}$$

Combined daily infection probability, per phase (applied sequentially as susceptibility is re-evaluated after each phase):

$$\pi_i(t) = 1 - \exp\Big(-\phi_{R(i)}\,\sigma_{n_i(t)}\,\Big[\lambda_i^{\text{comm}}(t) \text{ or } \beta_{HH}\lambda_i^{HH,1}(t) \text{ or } \beta_{DC}\lambda_i^{DC}(t) \text{ or } \beta_{HH}\lambda_i^{HH,2}(t)\Big]\Big). \tag{14}$$

### S6. Protection metrics and bootstrap inference

Attack rate across simulation replicates $r = 1, \ldots, R$ at coverage $c$, group $g$:

$$AR_g(c) = \frac{\sum_r cases_{g,r}(c)}{\sum_r n_{g,r}(c)}. \tag{15}$$

Bootstrap confidence intervals: for each of $B = 2{,}000$ iterations $b$, replicates are resampled with replacement, jointly (same resampled index set for both the immunized and unimmunized group) when estimating Direct, and independently for the numerator and denominator replicate sets when estimating Total and Indirect:

$$\widehat{\Delta}^{(b)} = 1 - \frac{AR^{*(b)}_{num}}{AR^{*(b)}_{den}}, \qquad CI_{95\%} = \Big[Q_{0.025}\Big(\widehat{\Delta}^{(1)}, \ldots, \widehat{\Delta}^{(B)}\Big),\ Q_{0.975}\Big(\widehat{\Delta}^{(1)}, \ldots, \widehat{\Delta}^{(B)}\Big)\Big]. \tag{16}$$

### S7. Calibration of monoclonal-antibody duration of immunity

The two mAb waning parameters used in the main model ($\omega^*_{mab}$ and $\omega_{mab}$; Table SS1) were calibrated with an age-structured compartmental model. The mAb follows a two-phase structure: an immunized infant remains fully protected for a mean of $D_{fix}$ days (Phase 1), then enters a partially protecting phase that wanes over a mean of $D_{mab}$ days (Phase 2). We fit $(D_{fix}, D_{mab})$ to the monthly vaccine-effectiveness (VE) estimates of Xu et al. [2026] against medically-attended RSV infection.

**Seasonal force of infection.** The infant cohort experiences a time-varying force of infection $\lambda(t)$ extracted from a burn-in of the full age-structured RSV transmission model (natural-history rates as in Section S1). For the infant age class,

$$\lambda(t) = \beta_0\left[1 + \text{Amp}\cos\left(\frac{2\pi\,(t - \phi P)}{P}\right)\right] \sum_{a'} \mathbf{C}_{a',\cdot} \times \frac{I_1(t) + \rho_1 I_2(t) + \rho_2\big(I_3(t) + I_4(t)\big) + (\text{vaccinated } I_k^V)}{N_{\text{tot}}(t)}, \tag{17}$$

with contact matrix $\mathbf{C}$, seasonal period $P = 52.18$ weeks, amplitude Amp, phase $\phi$. $\lambda(t)$ is evaluated over the 12-month post-introduction window and interpolated linearly for use in the cohort model below.

**Cross-sectional cohort design** VE at "month $m$ since receipt" is estimated from a distinct cohort that is already $(m-1)$ months post-dose and is then observed for exactly one month; VE is the incidence-density ratio over that window. Each cohort is described by a two-arm ODE (vaccinated with mAb; unvaccinated, same age) driven by the same $\lambda(t)$. Writing $\lambda \equiv \lambda(t)$, the *vaccinated* arm is

$$\frac{dM_b}{dt} = -\omega_{fix}\, M_b, \qquad \frac{dS_{ab}}{dt} = \omega_{fix}\, M_b - (\omega_{mab} + \lambda)\, S_{ab}, \tag{18}$$

$$\frac{dI_{ab}}{dt} = \lambda\, S_{ab} - \gamma_1\, I_{ab}, \qquad \frac{dMI_{ab}}{dt} = \gamma_1\, I_{ab} - \omega_{mab}\, MI_{ab}, \tag{19}$$

$$\frac{dS_0^V}{dt} = \omega_{mab}\, S_{ab} - \lambda\, S_0^V, \qquad \frac{dI_1^V}{dt} = \lambda\, S_0^V - \gamma_1\, I_1^V, \tag{20}$$

$$\frac{dS_1^V}{dt} = \gamma_1\, I_1^V + \omega_{mab}\, MI_{ab} - \sigma_1\lambda\, S_1^V, \qquad \frac{dI_2^V}{dt} = \sigma_1\lambda\, S_1^V - \gamma_2\, I_2^V, \tag{21}$$

$$\frac{dS_2^V}{dt} = \gamma_2\, I_2^V - \sigma_2\lambda\, S_2^V, \qquad \frac{dI_3^V}{dt} = \sigma_2\lambda\, S_2^V - \gamma_3\, I_3^V, \tag{22}$$

$$\frac{dS_3^V}{dt} = \gamma_3\, I_3^V + \gamma_3\, I_4^V - \sigma_3\lambda\, S_3^V, \qquad \frac{dI_4^V}{dt} = \sigma_3\lambda\, S_3^V - \gamma_3\, I_4^V, \tag{23}$$

and the *unvaccinated* arm is

$$\frac{dM}{dt} = -\omega_M\, M, \qquad \frac{dS_0}{dt} = \omega_M\, M - \lambda\, S_0, \tag{24}$$

$$\frac{dI_1}{dt} = \lambda\, S_0 - \gamma_1\, I_1, \qquad \frac{dS_1}{dt} = \gamma_1\, I_1 - \sigma_1\lambda\, S_1, \tag{25}$$

$$\frac{dI_2}{dt} = \sigma_1\lambda\, S_1 - \gamma_2\, I_2, \qquad \frac{dS_2}{dt} = \gamma_2\, I_2 - \sigma_2\lambda\, S_2, \tag{26}$$

$$\frac{dI_3}{dt} = \sigma_2\lambda\, S_2 - \gamma_3\, I_3, \qquad \frac{dS_3}{dt} = \gamma_3\, I_3 + \gamma_3\, I_4 - \sigma_3\lambda\, S_3, \tag{27}$$

$$\frac{dI_4}{dt} = \sigma_3\lambda\, S_3 - \gamma_3\, I_4. \tag{28}$$

Cumulative symptomatic incidence $C$ and person-time $T$ are integrated for each arm (the asymptomatic breakthrough $I_{ab}$ is excluded from $C$):

$$\frac{dC^V}{dt} = \lambda\, S_0^V + \sigma_1\lambda\, S_1^V + \sigma_2\lambda\, S_2^V + \sigma_3\lambda\, S_3^V, \qquad \frac{dC^U}{dt} = \lambda\, S_0 + \sigma_1\lambda\, S_1 + \sigma_2\lambda\, S_2 + \sigma_3\lambda\, S_3, \tag{29}$$

$$\frac{dT^V}{dt} = M_b + S_{ab} + I_{ab} + MI_{ab} + \sum_{k=0}^{3} S_k^V + \sum_{k=1}^{4} I_k^V, \quad \frac{dT^U}{dt} = M + \sum_{k=0}^{3} S_k + \sum_{k=1}^{4} I_k. \tag{30}$$

The model is run in weekly time steps, so a mean duration of $D$ days corresponds to a rate of $7/D$ per week: $\omega_{fix} = 7/D_{fix}$, $\omega_{mab} = 7/D_{mab}$, $\omega_M = 7/D_M$, and $\gamma_k = 7/d_k$ with $(d_1, d_2, d_3) = (10, 7, 5)$ days.

**Monthly VE and calibration.** For each target month $m$, the cohort is initialised at dosing ($M_b = 1$, $M = 1$, all other compartments and accumulators 0), integrated to the start of month $m$ (a pre-run of $(m-1)\,\bar{w}$ weeks, $\bar{w} = 4.345$ weeks per month, with the FOI clock offset so absolute time tracks the season), and then integrated for one further month. With $\Delta$ the increment of an accumulator over that one-month window,

$$VE(m) = 1 - \frac{\Delta C^V / \Delta T^V}{\Delta C^U / \Delta T^U}. \tag{31}$$

The durations are estimated by least squares against the monthly medians $VE^{obs}(m)$ of Xu et al.:

$$\big(\hat{D}_{fix}, \hat{D}_{mab}\big) = \arg \min_{D_{fix}, D_{mab}} \sum_{m=1}^{12} \big[ VE(m; D_{fix}, D_{mab}) - VE^{obs}(m) \big]^2, \tag{32}$$

minimised by a coarse grid search followed by L-BFGS-B refinement (bounds $D_{fix} \in [10, 200]$, $D_{mab} \in [10, 700]$ days). The fitted values define $\omega^*_{mab} = 7/\hat{D}_{fix}$ and $\omega_{mab} = 7/\hat{D}_{mab}$, which are carried into the main model (Table SS1).

### S8. Supplementary Tables

Table S1: **Default model parameters.** Household and daycare structures were constructed to reflect U.S. demographic characteristics (US Census Bureau [2023]), while the daycare structure was based on the observed structure of Phyllis Bodel Childcare Center [2026] and informed by Connecticut Office of Early Childhood [2026]. For the baseline transmission and force of infection parameters ($\beta_{DC}, \beta_{HH}, \lambda_{ext}$), we calibrate them to secondary attack rates for infants, toddlers and adults of RSV for different age groups from Hall et al. [1976] and Cohen et al. [2024]. See Fig. S17 in Supplementary Figures.

| **Parameter** | **Description** | **Value** | **Reference** |
|---|---|---|---|
| **Population and demography** | | | |
| `n_infants_dc` | Infants per centre | 90 | Assumed |
| `n_toddlers_dc` | Toddlers per centre | 90 | Assumed |
| `staff_child_ratio` | Staff-to-children ratio | 1:4 | Assumed |
| `n_parents` | Parents (adults) per household | 2 | Assumed |
| `p_two_caregiver` | Prob. household has two caregivers | 0.85 | Assumed |
| `p_elderly` | Elderly per household (0/1/2) | 0.92 / 0.07 / 0.01 | Assumed |
| `p_toddler` | Toddlers per household (0/1/2) | 0.70 / 0.25 / 0.05 | Assumed |
| `p_infant` | Infants per infant-household (0/1) | 0 / 1 (exactly one) | Assumed |
| `staff_hh_size` | Staff household size | 2 | Assumed |
| Infant age range | Age at study start (days) | 1–121 | Assumed |
| Toddler age range | Age at study start (days) | 366–1461 | Assumed |
| Adult age range | Age (days) | 9000–14000 | Assumed |
| Elderly age range | Age (days) | 23000–29000 | Assumed |
| Staff age | Fixed age (days) | 10000 | Assumed |
| **Daycare / classroom structure and contact** | | | |
| `cluster_size` | Classroom size | 9 | Assumed |
| $w_{in}$ | Within-classroom contact weight | 1.00 | Assumed |
| $w_{bw,child}$ | Between-classroom child weight | 0 | Assumed |
| $w_{bw,staff}$ | Between-classroom staff weight | 1 | Assumed |
| `daycare_entry_age` | Minimum age for daycare entry (days) | 120 | Assumed |
| `attendance_child` | Children attendance prob. | 1.00 | Assumed |
| `attendance_staff` | Staff attendance prob. | 1.00 | Assumed |
| `sick_attendance` | Symptomatic ($I$) attendance prob. | 0.3 | Shapiro [1984] |
| $C^{DC}$, $C^{HH}$ | Role-structured contact matrices | - | Litvinova et al. [2025] |
| **Natural history and immunity** | | | |

*continued on next page*

*Table S1 (continued)*

| Parameter | Description | Value | Reference |
|---|---|---|---|
| $\omega_M$ | Maternal-antibody waning rate | 1/112 $d^{-1}$ | Pitzer et al. [2015] |
| $\sigma_n$ | Rel. susceptibility after $n$ infections | 1, 0.76, 0.60, 0.40 ($n$=0, 1, 2, ≥3) | Pitzer et al. [2015] |
| $\rho_n$ | Rel. infectiousness by episode | 1, 0.75, 0.51 ($n$=1, 2, ≥3) | Pitzer et al. [2015] |
| $\mu(n)$ | Mean infectious duration (days) | 10 / 7 / 5 ($n$=1, 2, ≥3) | Pitzer et al. [2015] |
| $\phi_R$ | Role-specific susceptibility | infant 3.27; toddler 1.98; adult/elderly/staff 1.00 | Estimated |
| $\kappa_R$ | Role-specific infectiousness | infant 0.78; toddler 1.69; adult/elderly/staff 1.00 | Estimated |
| **Monoclonal antibody (Model A)** | | | |
| $\omega^*_{mab}$ | Full-protection phase waning ($M_b \to S_{ab}$; mean 53.6 d) | 1/53.6 $d^{-1}$ | Calibrated |
| $\omega_{mab}$ | Post-phase mAb waning ($S_{ab} \to S$, $MI_{ab} \to S$; mean 168.6 d) | 1/168.6 $d^{-1}$ | Calibrated |
| $\rho_{mab}$ | Rel. infectiousness of breakthrough infection ($I_{ab}$) | 0.20 | Munywoki et al. [2015] |
| `mab_age_max` | Maximum age at dosing (days) | 240 | Assumed |
| `rsv_season_months` | Dosing / season months | Oct–Mar | Assumed |
| **Viral load and infectiousness** | | | |
| $C_{peak}, r, d, t_{peak}$ | Ct trajectory (infant) | 33.72, 2.33, 0.53, 2.72 | Li et al. [2025] |
| $C_{peak}, r, d, t_{peak}$ | Ct trajectory (toddler) | 34.67, 1.79, 0.30, 2.94 | Li et al. [2025] |
| $C_{peak}, r, d, t_{peak}$ | Ct trajectory (adult/elderly/staff) | 34.34, 1.75, 0.19, 3.01 | Li et al. [2025] |
| $\mu_{vl}$ | Ct→infectiousness midpoint | 32.79 | Li et al. [2025] |
| $s_{vl}$ | Ct→infectiousness slope | 1.92 | Li et al. [2025] |
| $Ct_{LOD}$ | Ct limit of detection | 45 | Li et al. [2025] |
| $\beta_1$ | Baseline infectiousness intercept | 0.02 | Estimated |
| $\beta_2$ | Ct-dependent infectiousness scale | 0.02 | Estimated |
| **Transmission and force of infection** | | | |
| $\beta_{DC}$ | Daycare transmission scaling | 1.5 | Calibrated |
| $\beta_{HH}$ | Household transmission scaling | 1.5 | Calibrated |
| $\lambda_{ext}$ | Community importation baseline | $5 \times 10^{-4}$ $d^{-1}$ | Calibrated |
| **Severe-outcome risk** | | | |
| $p_{LRTI}$(infant) | P(LRTI \| infection), 1 / 2 / 3+ | 0.40 / 0.25 / 0.15 | Henderson et al. [1979], Ohuma et al. [2012], Glezen et al. [1986] |
| $p_{LRTI}$(toddler) | P(LRTI \| infection), order 1 / 2 / 3+ | 0.20 / 0.12 / 0.08 | Ohuma et al. [2012], Henderson et al. [1979] |

*continued on next page*

*Table S1 (continued)*

| **Parameter** | **Description** | **Value** | **Reference** |
|---|---|---|---|
| $p_{LRTI}$(adult) | P(LRTI \| infection), 1 / 2 / 3+ | 0.05 / 0.03 / 0.03 | Hall et al. [2001], Falsey and Walsh [2000] |
| $p_{LRTI}$(elderly) | P(LRTI \| infection), 1 / 2 / 3+ | 0.18 / 0.15 / 0.12 | Falsey et al. [2005], Falsey and Walsh [2000] |
| $p_{hosp}$(infant) | P(hospitalization \| infection), order 1 / 2 / 3+ | 0.030 / 0.015 / 0.008 | Nakajo and Nishiura [2022], Suss and Simões [2024] |
| $p_{hosp}$(toddler) | P(hospitalization \| infection), 1 / 2 / 3+ | 0.008 / 0.005 / 0.003 | Assumed Nakajo and Nishiura [2022] |
| $p_{hosp}$(adult) | P(hospitalization \| infection), 1 / 2 / 3+ | 0.001 / 0.001 / 0.001 | Hall et al. [2001], Volling et al. [2014], reinfection risk assumed |
| $p_{hosp}$(elderly) | P(hospitalization \| infection), 1 / 2 / 3+ | 0.050 / 0.050 / 0.050 | Falsey et al. [2005] reinfection risk assumed |

Table S2: **Detailed household composition.** Exact combinations of infants, toddlers, adults, and grandparents (elderly) per household, polled across all households in 50 simulated daycare centers.

| Infants | Toddlers | Adults | Elderly | n (%) |
|---|---|---|---|---|
| 1 | 0 | 2 | 0 | 2555 (36.5%) |
| 0 | 1 | 2 | 0 | 1548 (22.1%) |
| 1 | 1 | 2 | 0 | 843 (12.0%) |
| 0 | 2 | 2 | 0 | 422 (6.0%) |
| 1 | 0 | 1 | 0 | 409 (5.8%) |
| 0 | 1 | 1 | 0 | 286 (3.8%) |
| 1 | 0 | 2 | 1 | 177 (2.5%) |
| 1 | 1 | 1 | 0 | 170 (2.4%) |
| 1 | 2 | 2 | 0 | 148 (2.1%) |
| 0 | 1 | 2 | 1 | 120 (1.7%) |
| 0 | 2 | 1 | 0 | 65 (0.9%) |
| 1 | 1 | 2 | 1 | 55 (0.8%) |
| 1 | 0 | 1 | 1 | 33 (0.5%) |
| 1 | 0 | 2 | 2 | 32 (0.5%) |
| 0 | 2 | 2 | 1 | 29 (0.4%) |
| 1 | 2 | 1 | 0 | 27 (0.4%) |
| 0 | 1 | 1 | 1 | 19 (0.3%) |
| 0 | 1 | 2 | 2 | 18 (0.3%) |
| 1 | 2 | 2 | 1 | 13 (0.2%) |
| 1 | 0 | 1 | 2 | 12 (0.2%) |
| 1 | 1 | 2 | 2 | 12 (0.2%) |
| 1 | 1 | 1 | 1 | 11 (0.2%) |
| 0 | 2 | 1 | 1 | 5 (0.1%) |
| 0 | 2 | 2 | 2 | 5 (0.1%) |
| 1 | 2 | 1 | 1 | 2 (0.0%) |
| 0 | 1 | 1 | 2 | 2 (0.0%) |
| 0 | 2 | 1 | 2 | 1 (0.0%) |
| 1 | 2 | 2 | 2 | 1 (0.0%) |

Table S3: **Daycare structure under alternative assumptions (scenario 2).** Each simulated daycare center enrolled a fixed 90 infants (<1 year old) and 90 toddlers (1-4 years old), and children-staff ratio is fixed at 3. The remaining, variable structural characteristics are summarised below (mean [range] across 50 cohorts unless shown as n (%)).

| **Household structure** | **Value** |
|---|---|
| Households per centre | 90.0 (90.0–90.0) |
| Infant + toddler siblings, n (%) | 78.6 (87.3%) |
| Infant only, n (%) | 11.4 (12.7%) |
| Toddler only, n (%) | 0.0 (0.0%) |
| No child in home (e.g., grandparent-only), n (%) | 0.0 (0.0%) |
| Households with two caregivers, % | 84.6 (75.6–91.1) |
| Households with a grandparent, % | 7.6 (3.3–14.4) |

Hanmeng Xu, Camilla Aparicio-Llorente, Joshua Warren, Lee Kennedy-Shaffer, Virginia E. Pitzer, Daniel M. Weinberger, Carlos R. Oliveira, and PROVE-ID Group Authors. Durability and seasonal variation in the effectiveness of nirsevimab over three seasons in Connecticut. *medRxiv*, 2026. Preprint.
